# Impact of ‘Black Race’ coefficient in eGFR on Our Community and Medical Education

**DOI:** 10.1101/2022.05.25.22275472

**Authors:** Shibani Kanungo, Amy R. Lorber, Christine Schmitt, Kaitlyn VanRiper, Joseph Billian

## Abstract

A critical national conversation over the use of race in estimation of glomerular filtration rate (eGFR) was stimulated by medical students at the University of Washington over the past few years. They explored, with overdue scrutiny, the ways race is portrayed and used in medical education and practice. Questions were raised about the inclusion of the multiplicative factor for Black race in eGFR calculation, with implications for differential clinical management, and the possibility of disadvantaging patients, solely based on race. Their call for investigation and action, among the rising voices of many, started a reexamination of eGFR and the numerous other areas of medicine touched by racism. As a result, national groups of physicians, scientists, specialists, and patients (including the American Society of Nephrology and National Kidney Foundation) have called for removal of race from calculation of eGFR. We scrutinized use of ‘Black race’ coefficient in eGFR calculation and consequence of its use on our local community in SW Michigan. A cross-sectional analysis of electronic health record (EHR) data from local hospital systems demonstrates the impact removal of race from eGFR calculation could have on our community. Our work may serve as a prototype for ongoing and thorough examination of questions of race in medicine. Truly eradicating racism from medicine may require continuous critical evaluation of current established norms of medical education and care. As similar issues arise in medical education, we can identify how such changes shape healthcare on a local level and integrate this knowledge into our education.

**Key points:** *Question:* Can use of Black race in eGFR equation affects our local community?

*Findings:* Cross sectional study of a community population accessing local health care system for assessment of kidney function, found significant association between self-identified ‘Black’ population advance to CKD Stage 3 or higher upon removal of “black race” coefficient in eGFR MDRD calculation.

*Meaning:* Race use in medical practice and medical education further perpetuates systemic racism and widens health care access inequity.

## Introduction

### Racism in Medicine and Medical Education

As we seek to understand the use of race in medicine, it is necessary to revisit our history. The population of the United States (U.S.) is shaped by a history of migration from diverse locations around the globe along with the founding presence of Native Americans.^1^ The U.S. has never been a country of people from a homogenous origin, ancestry, or race. Incipient in our history, the forced migration of enslaved people from Africa was a critical in shaping the demographics of the U.S. The fact that slavery was a crucial driver of economic expansion of the early U.S. colonies created social need to justify the subjugation of enslaved peoples.^2^ Thus, even prior to the codification of U.S. as a country, the division of our population along culturally-defined racial groups was established. This “legacy of slavery” lives on, as a social hierarchy privileging White people above Black people – enforced by de jure and de facto means across centuries.^3^

Where there is subjugation, there is rationalization and attempts to explain or excuse oppression within biology, or ‘nature.’ Medicine and the study of human biology are at the center of this ongoing ‘justification.’ Attempts to embed racial superiority within ‘objective’ scientific knowledge is even present in the classification system for organisms put forth by Carl Linnaeus in System Naturae in 1758. He organized living beings from kingdom to species, but also further divided *Homo sapiens* into a biosocial hierarchy of “white, red, yellow, and black:” *europaeus, americanus, asiaticus*, and *afer*^4^ – placing ‘black race’ as most inferior. Shortly thereafter, the term “Caucasian” was coined, as referring to “white peoples of Europe.”^5^ This example demonstrates how our modern understanding of race is derived from defense of a racist hierarchy, whether explicitly or implicitly.^3^

Race is a social construct, rather than a biological one. Population studies consistently demonstrate significantly more variability in genetic markers between individuals of the same race than between racial categories overall.^6^ Yet the “one-drop” understanding of race, that any degree of Black ancestry means Black, was used a tool for segregation until the 1960s and still influences how people self-identify and understand racial identity.^7^ Even medical research attempting to capture “race” has been very inconsistent in providing opportunities to identify oneself as multiracial – reinforcing this binary concept of race in today’s constantly changing U.S. population demographics. Globalization, with 14% (46 million people) of the U.S. population born in another country, even further calls into question a homogenous concept of race.^7^

Our undergraduate medical education lays the foundation of our medical students’ biomedical knowledge and can critically, indoctrinate them with the values and culture of medicine. Medical education includes egregious instances of abuse, such as the Tuskegee Syphilis Study and the experimentation on enslaved women Lucy, Betsey, and Anarcha at the hands of Dr. J. Marion Sims.^8^ However, such horrific examples of abuse of the past gives students the perspective that contemporary medicine is free from institutionalized racism, but this is not the case. We assume contemporary medical education no longer promotes racial superiority, but it does still teach differences between racial groups. We often discuss the harms in the Tuskegee Syphilis study as solely the withholding of treatment from the study’s Black participants without their consent. However, the study itself was a recapitulation; a study of the long-term effects of syphilis had already been performed on White patients.^9^ This second study was only performed due to an underlying belief that there would be a difference in disease course in Black patients. Here we see the harm that arises when assumptions that race is a useful proxy for biology go unexamined.

Medical education teaches that race is as a “risk factor,” often used as a proxy for other factors which influence health.^2^ When race or racial differences in disease prevalence are discussed, they are often given without context and perpetuates the concept of inherent biological differences. The Liaison Committee of Medical Education (LCME) established guidelines for “cultural competency” in medical education to address inequities in care and poor health outcomes across racial and ethnic groups.^10^ However, “cultural competency” does not seem to address racism. Undergraduate medical education rarely takes the time to deconstruct and define the complexities of race and its relationship to medicine. Medical students witness the misapplication of race move from lecture into practice. Yet they are rarely encouraged to ask how our medical definitions of race can affect patients and their lives in our local communities.

A topic which has received national attention over the past several years is the calculation of estimated glomerular filtration rate (eGFR). In this article, we will see how using a multiplicative factor for Black race in eGFR can affect our local patient community.

### Introduction to Kidney Function and Physiology

Glomerular filtration rate is the most widely available and accepted measurement for kidney function.^11,12^ An individual’s glomerular filtration rate is the sum of the filtration of all of the functional kidney units as plasma is filtered across the glomerular capillaries.^11,12^

The direct measurement of kidney function presents logistical barriers. For this reason, clinicians and researchers have long sought calculations which accurately estimate true, or measured, GFR. Serum creatinine is produced endogenously by the body and its measurement is now a low-cost and ubiquitous indicator for kidney function.^13,14^ It is used because creatinine clearance is only ∼5-10% above that of inulin, an exogenous substance of which is a gold standard for accurately measuring GFR.^15,13,16^ The value of creatinine must be modified by an equation to estimate GFR, complicated by the variability of creatinine production, intake, and excretion, which has resulted in the inclusion of race, weight, sex, and age in calculation for a given individual.

The Modification of Diet in Renal Disease (MDRD) and, later, the Chronic Kidney Disease Epidemiology Collaboration (CKD-EPI) equations were developed in this way and are considered to be the best measures of eGFR by organizations such as the National Kidney Foundation (NKF) and the National Institute of Diabetes and Digestive and Kidney Diseases (NIDDK).^17^ Within the distribution of the United States, Non-Hispanic Black people have the highest rate of chronic kidney disease – at 16%. In the past, defense of the use of race in MDRD has cited the fact Black populations have a higher rate of chronic kidney disease as justification.^18^

Alternatives to the MDRD, CKD-EPI, and even use of serum creatinine (SCr) have been proposed. Cystatin C could be an alternate marker for eGFR and may be impacted by factors including gender, smoking status, height and weight, muscle mass, age, and CRP values, but perhaps to a lesser extent than creatinine ^19,20,21^ Cystatin C may also be a better predictor of all-cause mortality in patients.^22,23^ An amendment of the original, there exists a CKD-EPI-Cystatin C equation (2012) which only includes a correction for females and does not use a correction for race. The practical use of cystatin C may be limited due to laboratory diagnostic feasibility and longer turnaround time.^24,25^ Supplemental Table S1 gives a snapshot of equations used for eGFR.

### Staging of Chronic Kidney Disease and Implications for Treatment

Chronic Kidney Disease (CKD) is defined as damage to kidney morphology or function for at least 3 months and decreased kidney function as defined by eGFR as noted in Supplemental Table S1.^26,27,28^ CKD stages are defined using the Kidney Disease Improving Global Outcomes (KDIGO) clinical practice guidelines^29^ and staging influences healthcare outcomes by placing patients in different treatment categories for medication management, timing of referral to a nephrologist for specialty care, and being considered for kidney transplant (Table 1).

**Table 1:** CKD Staging.

| **CKD Stage** | **GFR (mL/min/1.73m<sup>2</sup>)<sup>35</sup>** | **General Management<sup>28</sup>** | **Additional Impacts<sup>36</sup>** |
| --- | --- | --- | --- |
| Stage 1 | ≥ 90 | Manage medically and **address comorbid conditions** | **Impacts drug dosing** (e.g. Use of SGLT-2 inhibitors in Type 2 DM limited to eGFR ≥ 30)<br>**Changes clinical interventions and screening** for related conditions (anemia, malnutrition, mineral and bone disorders)<br>Affects the results specified by **Kidney Failure Risk Calculator** |
| Stage 2 | 60 - 89 | **Track progression** and continue previous management |  |
| Stage 3 | 30 - 59 | **Address complications** and continue previous management |  |
| Stage 4 | 15 - 29 | **Nephrology consult indicated** (evaluation for renal replacement therapy and/or transplant) and continue previous management |  |
| Stage 5 (ESRD<sup>a</sup>) | < 15 | **Begin renal replacement therapy and/or transplant** |  |

Definitions of the Stages of Kidney disease according to the Center for Disease Control.<sup>35</sup>
Standard of management by stage according to the American Academy of Family
Physicians.<sup>28,36,37,38</sup> <sup>a</sup>ESRD is End-Stage Renal Disease and is another term used for Stage 5
CKD.

### Epidemiology of Race and Kidney Disease

To examine the impact of changing eGFR equations, we must first understand two things: (1) the populations identifying as White, Black, mixed race, or other and (2) the burden of kidney disease in these groups. The current epidemiology of race and kidney disease in the U.S. and in SW Michigan (Kalamazoo County) is detailed in Table 2.

**Table 2:**
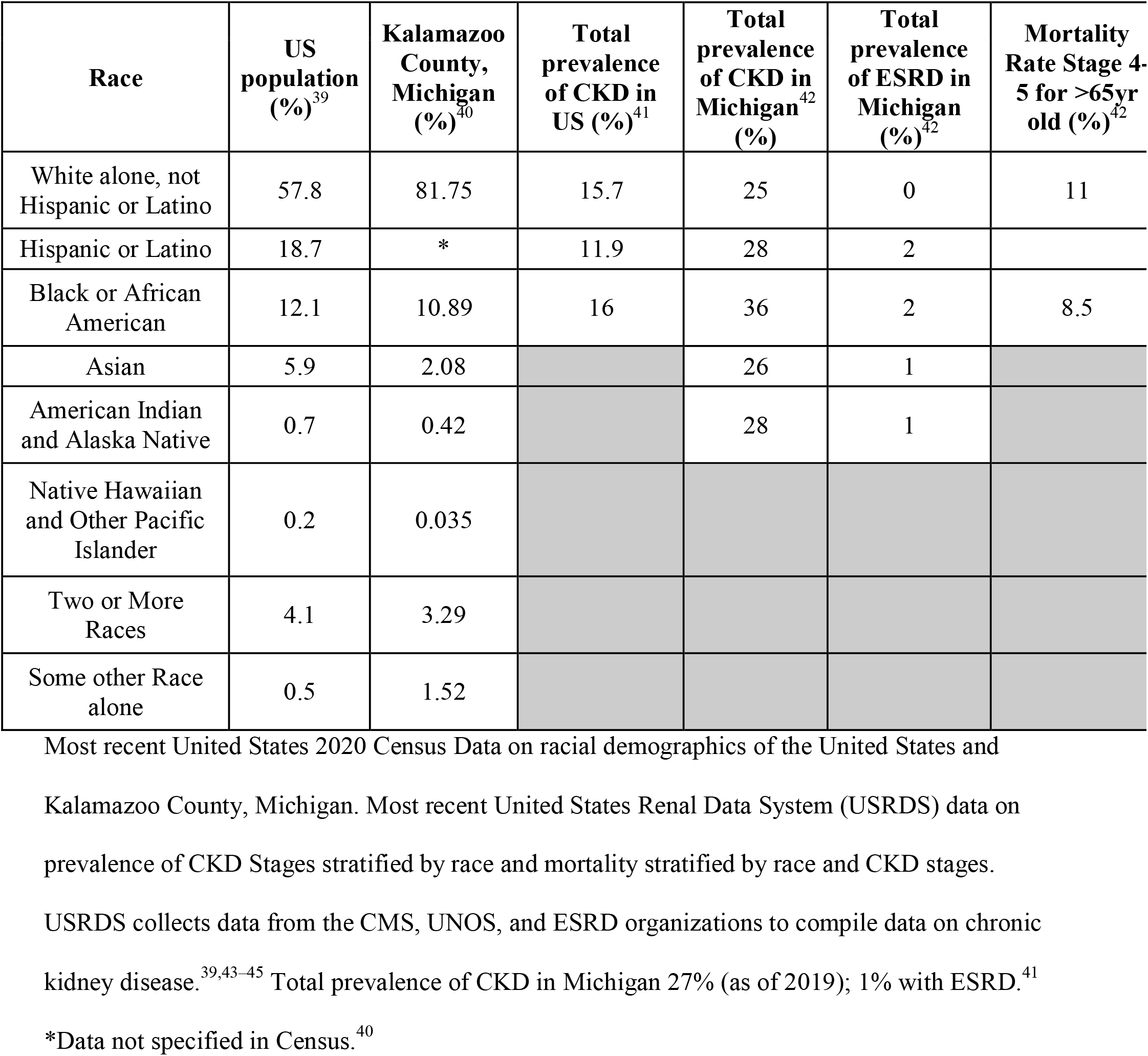
Demographics and Prevalence of CKD in U.S. and Michigan.

Justification of using Black race in determining kidney function in the MDRD equation is inconclusive and more research to assess the impact of removing race in eGFR is needed,^6^ especially since race biases could be associated with decisions like potential kidney transplant eligibility.^30^ In this paper, we have presented the broader context of race within medicine and the history eGFR calculations. We further investigated how the use of Black race in eGFR affects our local community in Southwest Michigan.

## Methods

We conducted a retrospective cross-sectional analysis to determine the burden of chronic kidney disease via eGFR and its variations between racial groups in Southwest Michigan. Nonidentifiable EHR data was collected from 1/1/2019 to 12/31/2019 as part of routine outpatient primary care visits at two local healthcare systems and a local outpatient clinic of an academic center. Variables of interest included age, race, gender, serum creatinine levels and calculated eGFRs (if any), zip code, ICD-10 code associated with the lab order, and visit type from which the order was placed and this information was provided by data brokers at each institution. For eGFR calculation, each location used the MDRD equation for each racial group and we then utilized KDIGO guidelines for CKD staging (Table 1).

Further analysis of eGFR calculation omitting the ‘Black race’ coefficient was determined for all subjects who self-identified as ‘Black’ in EHR. Prevalence of CKD and geographic distribution were assessed utilizing zip code. For patients with multiple serum creatinine measures within the study period, the highest value was taken into consideration in our results.

Inclusion Criteria included:

– age greater than or equal to 18 years
– accessing primary care services at outpatient clinics noted above
– serum creatinine testing from 1/1/2019 to 12/31/2019.

This study underwent review and was approved by Western Michigan University Homer Stryker MD School of Medicine IRB (IRB#:WMed-2020-0661).

### Statistical Methods

Data analysis was done utilizing statistical analysis software, SAS^®^ V9.4 (2013). eGFR was calculated from SCr, sex, race, and age according to the MDRD equation as noted in Supplemental Table S1. eGFR was categorized into CKD stages noted in Table 1. Patients’ ages were categorized into the following groups: 18-19; 20-29; 30-39; 40-49; 50-59; 60-69; 70+.

The primary objective was met with three chi-square tests for association. The overall type-1 error rate was controlled at a level of 5% using a Bonferroni-adjusted significance threshold (α) of 0.016.

A Wald-approximation 95% confidence interval was used to estimate the binomial proportion of Black patients whose CKD stage as determined by MDRD equation would advance if ‘Black race’ coefficient (*1.212) was omitted.

## Results

EGFR and associated CKD stage were calculated for all 131,863 patients. Characteristics of our study population is as noted in Supplemental Table S3, including CKD stages, demographic variables, and associated conditions.

The primary objective was met with three chi-square tests answering three different questions regarding use of MDRD equation in determining burden of CKD stages as detailed below. Our data provided an affirmative answer to all three questions as noted in Table 4 below.

**Table 3:** CKD stages by Race.

| **Race** | **Study population (%)** | **CKD Stage 1 (%)** | **CKD Stage 2 (%)** | **CKD Stage 3 (%)** | **CKD Stage 4 (%)** | **CKD Stage 5 (%)** |
| --- | --- | --- | --- | --- | --- | --- |
| All | 100% | 22% | 58% | 19% | 1.1% | 0.31% |
| White Alone | 86% | 19% | 60% | 20% | 1.1% | 0.24% |
| Black or African American alone | 7.8% | 41% | 45% | 12% | 1.3% | 0.93% |
| American Indian and Alaska Native alone | 0.42% | 23% | 57% | 18% | 1.3% | 0.72% |
| Asian alone | 1.4% | 40% | 52% | 6.8% | 0.44% | 0.39% |
| Native Hawaiian and other Pacific Islander alone | 0.09% | 27% | 55% | 16% | 0 | 1.8% |
| Some other race alone | 2.8% | 44% | 47% | 8.1%) | 0.94% | 0.52% |
| Two or more races | N/A | N/A | N/A | N/A | N/A | N/A |

**Table 4:** Prevalence of CKD Stages determined by MDRD Equation.

| **Prevalence of CKD Stages determined by MDRD Equation** |  |  |  |
| --- | --- | --- | --- |
|  | **Non-Black**<br>Frequency (Percent) | **Black**<br>Frequency (Percent) | Chi Square<br>**P-value** |
| Stages 3, 4 &<br>5 | 24,893 (20%) | 1,463 (14%) | < 0.0001 |
| Stages 4 & 5 | 1,593 (1.3%) | 229 (2.2%) | < 0.0001 |
| Stage 5 | 314 (0.26%) | 112 (1.10%) | < 0.0001 |
| **Total**<br>**Patients** | 121,643 | 10,220 |  |

Question 1: Is the rate of CKD Stage 3 or higher using MDRD equation different between Black and non-Black patients?

We found significant association between ‘Black’ and ‘not Black’ for CKD Stage 3 or higher with prevalence being 30% lower amongst Black (P-value < 0.0001.)

Question 2: Is the rate of Stage 4 or higher CKD different between Black and non-Black patients?

We found significant association between ‘Black’ and ‘not Black’ for CKD Stage 4 or higher with prevalence being 69% higher amongst Black patients (P-value < 0.0001.).

Question 3: Is the rate of Stage 5 CKD different between Black and non-Black patients?

We found significant association between the variable indicating ‘Black’ and ‘not Black’ for CKD Stage 5, with prevalence being more than 3 times higher amongst Black patients (P-value < 0.0001).

We determined that omission Black race coefficient (*1.212) in the MDRD equation would not impact any other race except subjects identifying themselves as Black race only. We found that no data would change for the non-Black patients, but the calculated eGFR of Black patients would reduce by 17.5%. The changes in CKD staging, as it would affect our study population self-identifying as only ‘Black,’ are noted in Table 5a below.

**Table 5a:** Advance in CKD Stage without MDRD ‘Black race’ Coefficient.

| CKD Stage (EGFR range) | Total Number of Black Patients, using MDRD Equation | Number (Percent) of Black Patients who would <u>ADVANCE</u> to next CKD stage using MDRD equation without ‘Black race’ coefficient |
| --- | --- | --- |
| Stage 1 ($\geq 90$) | 4147 | 2602 (63%) |
| Stage 2 (60 – 89) | 4610 | 1571 (34%) |
| Stage 3 (30 – 59) | 1234 | 109 (9%) |
| Stage 4 (15 – 29) | 117 | 0 |
| Stage 5 ($<15$) | 112 | N/A |
| TOTAL | 10220 | 4282 (42%) |

In our data, the 95% confidence interval for the proportion of Black patients who would advance to the next stage of CKD upon ignoring ‘Black race’ (using Wald-approximated Confidence Interval for binomial proportion) is between 41.1% and 43.0%. Since no patients moved from Stage 4 into Stage 5, we can say with certainty that the third chi-square test (Table 4) would not change and while the significance of the first two tests would not change, the results do change substantially. These changes are summarized in Table 5b.

**Table 5b:** Evaluating Changes in CKD Stage 3+, 4+, 5+.

|  |  | Using MDRD equation with ‘Black race’ coefficient |  | Calculating EGFR without ‘Black race’ coefficient |  |
| --- | --- | --- | --- | --- | --- |
|  | Prevalence, non-Black | Prevalence, Black | Difference between Black & non-Black (% of non-Black) | Prevalence, Black | Difference between Black & non-Black (% of non-Black) |
| **CKD Stage 3+** | 20% | 14% | -30% | 29% | +45% |
| **CKD Stage 4+** | 1.3% | 2.2% | +69% | 3.3% | +154% |
| **CKD Stage 5** | 0.26% | 1.10% | +323% | 1.10% | +323% |

For the first test of association between race and CKD Stage 3 or higher, the difference in disease prevalence would reverse. Using MDRD equation with the ‘Black race’ coefficient, the rate of CKD Stage 3 or higher was <u>30% lower</u> among Black patients than non-Black patients, but omitting the ‘Black race’ coefficient, the rate of CKD Stage 3+ would be <u>45% higher</u> among Black patients than non-Black patients.

For the second test of association between race and CKD Stage 4 or higher, the difference in disease prevalence would become more pronounced since the rate of CKD Stage 4+ was <u>69% higher</u> among Black patients than non-Black patients even using the ‘Black race’ coefficient. Omitting the ‘Black race’ coefficient, the rate of CKD Stage 4+ would increase to be <u>154% higher</u> among Black patients than non-Black patients.

There were no significant differences noted in demographic or associated condition characteristics amongst the Black patients who would advance a stage in CKD (n=4229) with those who would not advance (n=5921) (Supplemental Table S4).

## Discussion

During the data collection period in 2019, both hospital systems and the outpatient clinic site were all using MDRD.

Based on our data, 34% of Black patients in southwest Michigan with CKD would advance to Stage 3 from Stage 2 if the MDRD equation was used without ‘Black race’ coefficient. There are significant differences in management and treatment of CKD that occur when a patient enters Stage 3 CKD, including standard of referral to establish care with a nephrologist.^29^

Our data also finds that, with removal of the Black race coefficient, 9% of Black patients would advance to Stage 4 (from Stage 3) CKD. This is important because patients with a history of progressive CKD and an eGFR of ≤ 20 mL/min/1.73m^2^ (within Stage 4) qualify for addition to the renal transplant list.^29^

There is a disconnect between the proportions of Black vs non-Black patients with ESRD, as 66.6% more Black patients fall into Stage 5 when compared to non-Black patients (Supplemental Table S2) across the U.S. population. It may not be immediately apparent why there is an isolated, higher proportion of Black patients in ESRD, given the lower rates for this patient group across the first four CKD stages when compared to non-Black patients. In fact, there are mortality benefits to early referral to a nephrologist in the first stages of CKD.^31^ The eGFR calculations which place Black patients in lower CKD stages initially may deprive them of important treatment and referral early in their disease course. As early escalation in care slows or prevents the progression to ESRD, it appears that Black patients are systematically excluded from this benefit, and it seems reasonable to infer this may contribute to the increased proportion of Black patients who progress to Stage 5 CKD.

Many laboratories report CKD Stages 1 and 2 as “>60,” and only enumerate eGFR values, and therefore discrete staging, below 60 mL/min/1.73m^2^ – beginning with CKD Stage 3. Our data finds that of Black patients in our community currently in Stage 2, whose data may currently be reported as “>60,” and of no concern, would be moved into Stage 3 by removal of the race coefficient from eGFR calculation. This has significant implications for provider awareness and disease management.

Removal of the Black race coefficient allows for referral to a nephrologist, Medicare coverage, and potentially need for transplant and/or dialysis. CKD is also an independent risk factor for other comorbid conditions, such as cardiovascular disease. Earlier CKD diagnosis could allow providers to manage risk factors preemptively, like tightening diabetic, hypertensive, and lipid control, and potentially affect Medicare coverage for individuals who need transplant or dialysis, alleviating financial burden of ESRD.^32,33,29^ While it is considered on a case-by-case basis within Stage 5, expediting transplant eligibility could help some patients financially.

Our analysis of eGFR calculation in our community of SW Michigan establishes that a significant number of Black patients would be advanced to CKD Stage 2 (from 1), and from Stage 3 (from 2) with the removal of the race coefficient. Given the call for better detection, earlier awareness, and more prompt referrals, removal of the race coefficient would clearly move us toward those goals in our own community.

### Thoughts on Future Directions

In 2021, the American Society of Nephrology (ASN) and the National Kidney Foundation (NKF) published a recommendation on removal of race from eGFR and use of the CKD-EPI equation refit without race, which is now in use on their website (Supplemental Table S1).^34^ Considering these updated recommendations along with the results of our work, there are several future directions for investigation, including:

– Analysis of long-term patient outcomes as the removal of ‘Black race’ coefficient is implemented
– Evaluation of the updated eGFR calculation on pediatric, elderly, and other population groups with comorbid conditions (chronic illness, immunosuppression, etc.)
– Follow-up on how and when this new recommendation and underlying reasoning are integrated into medical education and medical student literature (textbooks, didactic materials, question banks).

We expect robust research on these topics to continue and hope that medical students will remain unafraid to question the status quo and move medicine forward.

## Supporting information

Impact eGFR Supplement

## Data Availability

All data produced in the present study are available upon reasonable request to the authors

## Acknowledgements

Jeffrey P Pearson, MD (Pathology) for discussion on eGFR calculations during study planning period; Ashutosh Goel, MD (Chief Medical Information Officer) for discussions on EHR repository and during study planning period; EHR data brokers, Rebecca Reardon (medical student) for IRB protocol development.

## Contribution

Medical students (Amy R. Lorber, Christine Schmitt, Kaitlyn VanRiper) – equal contribution in writing of manuscript ‘Introduction’.

Biostatistician (Joseph Billian) – Statistical Analysis, Tables, Maps

Faculty (Shibani Kanungo) – Concept, Design, Method, writing of complete manuscript, critical review for important intellectual content

## Ethical Approval

**(IRB #)** Western Michigan University Homer Stryker MD School of Medicine (IRB#:WMed-2020-0661).

## Disclaimer

None

This work has not been previously published and is not under consideration in the same or substantially similar form in any other journal.

## Bibliography

1. Dai CL, Vazifeh MM, Yeang C-H, et al. Population Histories of the United States Revealed through Fine-Scale Migration and Haplotype Analysis. Am J Hum Genet. 2020;106(106):371–388. doi:10.1016/J.AJHG.2020.02.002

2. Nieblas-Bedolla E, Christophers B, Nkinsi NT, Schumann PD, Stein E. Changing How Race Is Portrayed in Medical Education: Recommendations from Medical Students. Acad Med. 2020;95(95):1802–1806. doi:10.1097/ACM.0000000000003496

3. O’Connell HA, Curtis KJ, Dewaard J. Population change and the legacy of slavery. Soc Sci Res. 2020;87:102413. doi:10.1016/j.ssresearch.2020.102413

4. Mohsen H. Focus: Skin: Race and Genetics: Somber History, Troubled Present. Yale J Biol Med. 2020;93(93):215. Accessed August 24, 2021. /pmc/articles/PMC7087058/

5. Roberts D. Fatal Invention: How Science, Politics, and Big Business Re-Create Race in the Twenty-First Century. New York and London: The New Press; 2011.

6. Eneanya ND, Yang W, Reese PP. Reconsidering the Consequences of Using Race to Estimate Kidney Function. JAMA - J Am Med Assoc. 2019;322(322):113–114. doi:10.1001/jama.2019.5774

7. Adhikari K, Camilo Chacón-Duque J, Mendoza-Revilla J, Fuentes-Guajardo M, Ruiz-Linares A. The Genetic Diversity of the Americas. Annu Rev Genom Hum Genet. 2017;18:277–296. doi:10.1146/annurev-genom-083115-022331

8. Wailoo, Keith, (Department of History PU. The use of blacks for medical experimentation and demonstration in the old South. JAMA - J Am Med Assoc. 2018;320(320):1531–1533. doi:10.2307/2207450

9. Brandt AM. Racism and Research: The Case of the Tuskegee Syphilis Study. Hastings Cent Rep. 1978;8(8):21. doi:10.2307/3561468

10. Vasquez Guzman CE, Sussman AL, Kano M, Getrich CM, Williams RL. A Comparative Case Study Analysis of Cultural Competence Training at 15 U.S. Medical Schools. Acad Med. 2021;96(96):894–899. doi:10.1097/ACM.0000000000004015

11. Pollak MR, Quaggin SE, Hoenig MP, Dworkin LD. The Glomerulus: The Sphere of Influence. Clin J Am Soc Nephrol. 2014;9(9):1461. doi:10.2215/CJN.09400913

12. Levey AS, Becker C, Inker LA. Glomerular Filtration Rate and Albuminuria for Detection and Staging of Acute and Chronic Kidney Disease in Adults: A Systematic Review. JAMA. 2015;313(313):837. doi:10.1001/JAMA.2015.0602

13. Kashani K, Rosner MH, Ostermann M. Creatinine: From physiology to clinical application. Eur J Intern Med. 2020;72(October 2019):9–14. doi:10.1016/j.ejim.2019.10.025

14. Perrone RD, Madias NE, Levey AS. Serum Creatinine as an Index of Renal Function: New Insights into Old Concepts. Vol 38.; 1992. Accessed May 17, 2021. https://academic.oup.com/clinchem/article/38/10/1933/5650028

15. Pasala S, Carmody JB. How to use… serum creatinine, cystatin C and GFR. Arch Dis Child Educ Pract Ed. 2017;102(102):37–43. doi:10.1136/archdischild-2016-311062

16. Levey AS, Coresh J, Greene T, et al. Using standardized serum creatinine values in the modification of diet in renal disease study equation for estimating glomerular filtration rate. Ann Intern Med. 2006;145(145):247–254. doi:10.7326/0003-4819-145-4-200608150-00004

17. eGFR Calculator | National Kidney Foundation. Accessed October 14, 2021. https://www.kidney.org/professionals/kdoqi/gfr_calculator

18. Levey AS, Bosch JP, Lewis JB, Greene T, Rogers N, Roth D. Annals of Internal Medicine A More Accurate Method To Estimate Glomerular Filtration Rate from Serum Creatinine: A New Prediction Equation. Vol 130.; 1999. Accessed May 22, 2021. http://www.acponline.org.

19. Ferguson TW, Komenda P, Tangri N. Cystatin C as a biomarker for estimating glomerular filtration rate. Curr Opin Nephrol Hypertens. 2015;24(24):295–300. doi:10.1097/MNH.0000000000000115

20. Knight EL, Verhave JC, Spiegelman D, et al. Factors Influencing Serum Cystatin C Levels Other than Renal Function and the Impact on Renal Function Measurement. Vol 65.; 2004. doi:10.1111/j.1523-1755.2004.00517.x

21. Inker LA, Schmid CH, Tighiouart H, et al. Estimating Glomerular Filtration Rate from Serum Creatinine and Cystatin C. N Engl J Med. 2012;367(367):20–29. doi:10.1056/nejmoa1114248

22. Levey AS, Inker LA, Coresh J. GFR estimation: From physiology to public health. Am J Kidney Dis. 2014;63(63):820–834. doi:10.1053/j.ajkd.2013.12.006

23. Astor BC, Levey AS, Stevens LA, Van Lente F, Selvin E, Coresh J. Method of glomerular filtration rate estimation affects prediction of mortality risk. J Am Soc Nephrol. 2009;20(20):2214–2222. doi:10.1681/ASN.2008090980

24. Lewis J, Greene T, Appel L, et al. A comparison of iothalamate-GFR and serum creatinine-based outcomes: Acceleration in the rate of GFR decline in the African American Study of Kidney Disease and Hypertension. J Am Soc Nephrol. 2004;15(15):3175–3183. doi:10.1097/01.ASN.0000146688.74084.A3

25. Diao JA, Inker LA, Levey AS, Tighiouart H, Powe NR, Manrai AK. In Search of a Better Equation — Performance and Equity in Estimates of Kidney Function. N Engl J Med. 2021;384(384):396–399. doi:10.1056/nejmp2028243

26. What Is Chronic Kidney Disease? | NIDDK. Accessed September 2, 2021. https://www.niddk.nih.gov/health-information/kidney-disease/chronic-kidney-disease-ckd/what-is-chronic-kidney-disease

27. Versino E, Piccoli GB. Chronic kidney disease: The complex history of the organization of long-term care and bioethics. why now, more than ever, action is needed. Int J Environ Res Public Health. 2019;16(5). doi:10.3390/ijerph16050785

28. Johnson CA, Levey AS, Coresh J, Levin A, LauEknoyan JG. Clinical Practice Guidelines for Chronic Kidney Disease in Adults: Part 1. Definition, Disease Stages, Evaluation, Treatment, and Risk Factors. Am Fam Physician. 2004;70(70):869–876. Accessed August 15, 2021. http://www.aafp.org/afp.

29. KDIGO 2012 Clinical Practice Guideline for the Evaluation and Management of Chronic Kidney Disease. Kidney Int. 2013;3(1).

30. Zelnick LR, Leca N, Young B, Bansal N. Association of the Estimated Glomerular Filtration Rate with vs without a Coefficient for Race with Time to Eligibility for Kidney Transplant. JAMA Netw Open. 2021;4(4):e2034004–e2034004. doi:10.1001/jamanetworkopen.2020.34004

31. Winkelmayer WC, Owen WF, Levin R, Avorn J. A Propensity Analysis of Late Versus Early Nephrologist Referral and Mortality on Dialysis. J Am Soc Nephrol. 2003;14:486–492. doi:10.1097/01.ASN.0000046047.66958.C3

32. de Boer IH, Caramori ML, Chan JCN, et al. Executive summary of the 2020 KDIGO Diabetes Management in CKD Guideline: evidence-based advances in monitoring and treatment. Kidney Int. 2020;98(98):839–848. doi:10.1016/J.KINT.2020.06.024

33. Ronco P, Rovin B, Schlöndorff D, et al. KDIGO Clinical Practice Guideline for The Evaluation and Management of Chronic Kidney Disease. 2021;99(3). https://www.elsevier.com/books-and-journals/

34. Delgado C, Baweja M, Crews DC, et al. A Unifying Approach for GFR Estimation: Recommendations of the NKF-ASN Task Force on Reassessing the Inclusion of Race in Diagnosing Kidney Disease. Am J Kidney Dis. 2021;0(0). doi:10.1053/J.AJKD.2021.08.003

35. CDC - Chronic Kidney Disease - FAQ. Accessed September 2, 2021. https://nccd.cdc.gov/ckd/help.aspx?section=F#3

36. Gaitonde DY, Cook DL, Rivera IM. Chronic Kidney Disease: Detection and Evaluation. Am Fam Physician. 2017;96(96):776–783. Accessed October 14, 2021. http://www.aafp.org/afp.

37. Arnold MJ, Buelt A. Chronic Kidney Disease: Evaluation and Treatment Guidelines from the VA/DoD. Am Fam Physician. 2020;102(102):378–379.

38. Tangri N, Stevens LA, Griffith J, et al. A Predictive Model for Progression of Chronic Kidney Disease to Kidney Failure. JAMA. 2011;305(305):1553–1559. doi:10.1001/JAMA.2011.451

39. Bureau UC. Racial and Ethnic Diversity in the United States: 2010 Census and 2020 Census. Accessed September 2, 2021. https://www.census.gov/library/visualizations/interactive/racial-and-ethnic-diversity-in-the-united-states-2010-and-2020-census.html

40. Census - Table Results. Accessed September 2, 2021. https://data.census.gov/cedsci/table?q=Kalamazoo County Race and Ethnicity&tid=DECENNIALPL2010.P1

41. Annual Data Report | USRDS. Accessed October 15, 2021. https://adr.usrds.org/2020/chronic-kidney-disease/1-ckd-in-the-general-population).

42. End-Stage Renal Disease (ESRD) | CMS. Accessed September 6, 2021. https://www.cms.gov/Medicare/Coordination-of-Benefits-and-Recovery/Coordination-of-Benefits-and-Recovery-Overview/End-Stage-Renal-Disease-ESRD/ESRD

43. Annual Data Report | USRDS. Accessed September 2, 2021. https://adr.usrds.org/2020/chronic-kidney-disease/1-ckd-in-the-general-population

44. USRDS. Chapter 3: Morbidity and Mortality in Patients With CKD.

45. Mapping Medicare Disparities by Population - Centers for Medicare & Medicaid Services Data. Accessed September 2, 2021. https://data.cms.gov/tools/mapping-medicare-disparities-by-population

46. Musso CG, Joaquín Álvarez-Gregori ·, Jauregui · José, Macías-Núñez JF. Glomerular filtration rate equations: a comprehensive review. Int Urol Nephrol. 2016;48:1105–1110. doi:10.1007/s11255-016-1276-1

47. Rule AD, Bergstralh EJ, Slezak JM, Bergert J, Larson TS. Glomerular filtration rate estimated by cystatin C among different clinical presentations. Kidney Int. 2006;69(69):399–405. doi:10.1038/SJ.KI.5000073

48. Poggio ED, Wang X, Greene T, Lente F Van, Hall PM. Performance of the Modification of Diet in Renal Disease and Cockcroft-Gault Equations in the Estimation of GFR in Health and in Chronic Kidney Disease. J Am Soc Nephrol. 2005;16(16):459–466. doi:10.1681/ASN.2004060447

49. Peake M, Whiting M. Measurement of Serum Creatinine – Current Status and Future Goals. Clin Biochem Rev. 2006;27(27):173. Accessed October 15, 2021. /pmc/articles/PMC1784008/

50. Levey AS, Stevens LA, Schmid CH, et al. A new equation to estimate glomerular filtration rate. Ann Intern Med. 2009;150(150):604–612. doi:10.7326/0003-4819-150-9-200905050-00006

51. Porrini E, Ruggenenti P, Luis-Lima S, et al. Estimated GFR: time for a critical appraisal. Nat Rev Nephrol. 2019;15(15):177–190. doi:10.1038/s41581-018-0080-9

