## Supplementary material for "Impact of ‘Black Race’ coefficient in eGFR on Our Community and Medical Education": Impact eGFR Supplement

**Supplemental Tables and Figures:**

**Table S1: Equations for eGFR**

| **Equation** | **Calculation** | **Possible Benefits** | **Possible Harms** |
| --- | --- | --- | --- |
| **Cockcroft Gault (1973)** | *C_Cr_ = {(140-age) x kg)/(72 x S_Cr_)}*    multiply by 0.85 for female gender | Established a standard formula to estimate GFR incorporating gender differences for increased accuracy.^46^ |  |
| **MDRD (2005)** | *eGFR = 175 x (S_Cr_)^-1.154^ x (age)^-0.203^*    multiply by 0.742 for female gender  multiply by 1.212 for Black race | MDRD has been shown to be a close estimate of true GFR in patients with CKD or a kidney transplant.^47^    MDRD may be a better predictor of eGFR than the Cockcroft-Gault equation in patients with CKD with a true GFR <60.^48^ | MDRD was generated in 1999 by study of patients with renal disease – notably excluding patients without kidney disease, diabetic patients, those younger than 18 or older than 70, pregnant women, or transplant recipients – and therefore may not be appropriate for these groups.^16^  MDRD eGFR may not be representative of the general population and has been shown to underestimate kidney function in healthy patients.^48^  The MDRD utilized a form of this reaction for measurement – a kinetic alkaline picrate assay – which introduces variability in results.^49^ |
| **CKD-EPI Creatinine (2009)** | *eGFR = 141 x min(S_Cr_/*𝝹*, 1)^ɑ^ x (max(S_Cr_/*𝝹*, 1)^-1.209^ x 0.993^age^*    multiply by 1.018 for female gender  multiply by 1.159 for Black race  κ is 0.7 for females and 0.9 for males  α is -0.329 for females and -0.411 for males  *min* indicates the minimum of Scr/κor 1  *max* indicates the maximum of Scr/κ or 1 | Developed from a metanalysis of multiple studies and further refined relative to MDRD.^50^ | Asian and Hispanic populations were not well represented by creatinine equation studies.^25^  Development of the equation included a limited number of individuals over 70 or those from non-black racial minorities other than.^50^ |
| **CKD-EPI Creatinine-Cystatin (2012)** | *eGFR = 135 x min(S_Cr_/*𝝹*, 1)^ɑ^ x (max(S_Cr_/*𝝹*, 1)^-0.601^ x min(S_cys_/0.8, 1)^-0.375^ x (max(S_cys_/0.8, 1)^-0.711^ x 0.995^Age^*    multiply by 0.969 for female gender  multiply by 1.08 for Black race | eGFR calculation combining cystatin C and creatinine is closer to measured GFR than is creatinine alone.^23^ | This equation requires the use of creatinine and a correction for patient’s race. |
| **CKD-EPI Cystatin C (2012)** | *eGFR = 133 x min(S_cys_/0.8, 1)^-0.499^ x (max(S_cys_/0.8, 1)^-1.328^ x 0.996^Age^*    multiply by 0.932 for female gender | Cystatin C may be less impacted by “social demographics” than is serum creatinine.^21^  Cystatin C has improved prediction for mortality relative to Cr.^23^  Modestly improved correlations to GFR in patients with CKD compared to the MDRD, Cockcroft-Gault, or serum creatinine equations.^47^ | Cystatin C, controversy over whether it’s correlated to any other diseases or physiological measures other than GFR.^19^ Cystatin C may be impacted by gender, smoking status, height, weight, age, and CRP.^20^    Limited assay availability and longer turnaround time for Cystatin C.^25^ |
| **CKD-EPI (2021)** | *eGFR = 142 x min(standardized S_cr_/*𝝹*, 1)^α^ x max(standardized S_cr_/*𝝹*, 1)^-1.200^ x 0.9938^Age^ x 1.012 [if female]*  eGFR (estimated glomerular filtration rate) = mL/min/ 1.73 m^2^  S_cr_ (serum creatinine) = mg/dL  𝝹 = 0.7 (females) or 0.9 (males)  α = -0.241 (females) or -0.302 (males)  min = indicates the minimum of S_cr_/𝝹 or 1  max = indicates the maximum of S_cr_/𝝹 or 1 | Developed recently through consensus of a national task force of the ASN-NKF through extensive metanalysis.^34^ | This equation has not yet been widely implemented and long-term clinical outcomes are unknown. |
| *Note regarding the imprecision of eGFR calculation:* The standard for acceptable GFR estimates has been widely accepted as the “P30,” which is defined as the percentage of eGFR values which fall within ± 30% of a measured GFR value. This threshold was set without clinical or statistical underpinning.^51^ Furthermore, the acceptable value for P30 for a given equation is not well established; the CKD-EPI and MDRD achieve P30 values of 84.1% vs. 80.6%, respectively.^50^ One meta-analysis of >70 studies involving approximately 40,000 patients demonstrated that modern eGFR calculations may assign the incorrect CKD stage in 30-60% of patients.^51^ | | | |

**Table S2: Prevalence of CKD Stages by Race**

| **Race** | **Population, U.S. (%)** | **CKD Stage** | | | | | **Total prevalence of CKD** |
| --- | --- | --- | --- | --- | --- | --- | --- |
|  |  | **1** | **2** | **3** | **4** | **5** |  |
| All |  | 4.7 | 3.3 | 6.4 | 0.4 | 0.1 | 14.9 |
| Non-Hispanic White | 72 | 3.8 | 3.5 | 8 | 0.3 | 0.1 | 15.7 |
| Non-Hispanic African American/Black | 12.8 | 5.9 | 3.7 | 5.4 | 0.7 | 0.3 | 16 |
| Hispanic/Latino |  | 6.8 | 2.1 | 2.6 | 0.3 | 0.1 | 11.9 |

Data from years 2015-2018. Most recent United States Renal Data System (USRDS) data on prevalence of CKD Stages stratified by race. USRDS collects data from the CMS, UNOS, and ESRD organizations to compile data on chronic kidney disease.^41^

**Table S3: Study Population Demographics**

|  |  | **Frequency (Percent)** |
| --- | --- | --- |
| CKD Stage | 1 | 29,117 (22.1%) |
|  | 2 | 76,391 (57.9%) |
|  | 3 | 24,533 (18.6%) |
|  | 4 | 1,413 (1.1%) |
|  | 5 | 409 (0.3%) |
| Gender | Female | 75,908 (57.6%) |
|  | Male | 55,945 (42.4%) |
|  | Other | *[not reported if ≤ 10]* |
| Age | 18-19 | 1,479 (1.1%) |
|  | 20-29 | 9,572 (7.3%) |
|  | 30-39 | 13,656 (10.4%) |
|  | 40-49 | 18,368 (13.9%) |
|  | 50-59 | 25,361 (19.2%) |
|  | 60-69 | 31,242 (23.7%) |
|  | 70+ | 32,185 (24.4%) |
| Race | Black | 10,220 (7.8%) |
|  | Not Black | 121,643 (92.3%) |
| Nephropathy | Yes | 6,366 (4.8%) |
|  | No | 125,497 (95.2%) |
| Diabetes | Yes | 17,069 (12.9%) |
|  | No | 114,794 (87.1%) |
| Hypertension | Yes | 30,610 (23.2%) |
|  | No | 101,253 (76.8%) |

**Table S4: Traits and Conditions in CKD-stage-non-advancing Black patients and CKD-stage-advancing Black patients**

| **Trait or Condition** | **Proportion among CKD-stage-non-advancing Black patients** | **Proportion among CKD-stage-advancing Black patients** |
| --- | --- | --- |
| Age = 18-19 | 2% | 1% |
| Age = 20-29 | 11% | 9% |
| Age = 30-39 | 11% | 14% |
| Age = 40-49 | 16% | 19% |
| Age = 50-59 | 20% | 21% |
| Age = 60-69 | 22% | 21% |
| Age = 70+ | 18% | 15% |
| Gender = Female | 60% | 59% |
| Nephropathy | 7% | 4% |
| Diabetes | 14% | 15% |
| Hypertension | 22% | 23% |
